# Quantifying the effects of non-pharmaceutical and pharmaceutical interventions against COVID-19 epidemic in the Republic of Korea: Mathematical model-based approach considering age groups and the Delta variant

**DOI:** 10.1101/2021.11.01.21265729

**Authors:** Youngsuk Ko, Victoria May P. Mendoza, Yubin Seo, Jacob Lee, Yeonju Kim, Donghyok Kwon, Eunok Jung

## Abstract

**Background:** Early vaccination efforts and non-pharmaceutical interventions were insufficient to prevent a surge of coronavirus disease 2019 (COVID-19) cases triggered by the Delta variant. This study aims to understand how vaccination and variants contribute to the spread of COVID-19 so that appropriate measures are implemented.

**Methods:** A compartment model that includes age, vaccination, and infection with the Delta or non-Delta variants was developed. We estimated the transmission rates using maximum likelihood estimation and phase-dependent reduction effect of non-pharmaceutical interventions (NPIs) according to government policies from 26 February to 8 October 2021. We extended our model simulation until 31 December considering the initiation of eased NPIs. Furthermore, we also performed simulations to examine the effect of NPIs, arrival timing of Delta variant, and speed of vaccine administration.

**Results:** The estimated transmission rate matrices show distinct pattern, with the transmission rates of younger age groups (0 39 years) much larger than non-Delta. Social distancing (SD) level 2 and SD4 in Korea were associated with transmission reduction factors of 0.64 to 0.69 and 0.70 to 0.78, respectively. The easing of NPIs to a level comparable to SD2 should be initiated not earlier than 16 October to keep the number of severe cases below the capacity of Korea’s healthcare system. Simulation results also showed that a surge prompted by the spread of the Delta variant can be prevented if the number of people vaccinated daily was larger.

**Conclusions:** Simulations showed that the timing of easing and intensity of NPIs, vaccination speed, and screening measures are key factors in preventing another epidemic wave.

**Key Messages:**

- Maximum likelihood estimation can be utilized to determine the transmission rates of the Delta and non-Delta variants.
- The phase-dependent NPIs implemented by the Korean government were effectively quantified in the modelling study.
- Even with fast vaccination, resurgence of cases is still possible if NPIs are eased too early or screening measures are relaxed.
- The model can be used as a guide for policy makers on deciding appropriate SD level that considers not only disease control, but also the socio-economic impact of maintaining strict measures.

## 3 Introduction

The coronavirus disease 2019 (COVID-19) is an ongoing pandemic caused by the severe acute respiratory syndrome coronavirus 2 (SARS-CoV-2). The rise in the number of COVID-19 cases observed worldwide since July 2021 can be attributed to the emergence of the Delta variant of SARS-CoV-2 [1]. The Delta variant significantly reduced the effectiveness of vaccines to symptomatic infection to about 50-70% compared to the Alpha variant [2, 3]. Nevertheless, these vaccines are reported to still be effective against severe disease, hospitalization, and death [2, 4].

To mitigate the spread of COVID-19, on top of vaccination, various non-pharmaceutical interventions (NPIs), such as social distancing (SD), are still implemented concurrently. In Korea, a four-level SD plan has been implemented since July 2021. The plan outlines the number of persons allowed in a gathering and operational guidelines of shops, restaurants, gyms, pubs, concert halls, sports stadium, and other commercial facilities. Since 12 July 2021, SD level has been raised to the highest (SD4) to combat the fourth wave of COVID-19 in the Republic of Korea. As of 29 October 2021, there are 360 536 confirmed cases and 2817 deaths of COVID-19 in Korea [5].

Vaccination in Korea began on 26 February 2021 and priority was given to the healthcare workers and elderly. The vaccination plan proceeded according to age, starting with older age group [5]. Several types of vaccines have been administered, mostly BNT162b2 or ChAdOx1 [5]. Meanwhile, the first case of a local community transmission in Korea with the Delta variant was reported on 27 April 2021. The number of cases has steeply increased since then and the proportion of the Delta variant is almost 100% among the genome-sequenced COVID-19 cases in Korea [6].

Transmission rates between age groups before and during the period when Delta became the dominant variant of SARS-CoV-2 infections are estimated using MLE, which is a preferred parameter estimation and inference tool in statistics [7, 8]. The key factors leading the epidemic of COVID-19 recently are vaccination, variants of the virus, and implementation of SD. In this study, we developed an age-structured compartment model that captures COVID-19 transmission with the Delta and non-Delta variants among vaccinated and unvaccinated people. Furthermore, we quantify NPIs according to government policy and present scenarios that investigate the impact of timely vaccination, effective NPIs, and early detection of cases.

## 4 Materials and Methods

### 4.1 Data

We analyzed the COVID-19 case data including age, date of diagnosis by PCR test, and date of symptom onset from 26 February to 10 September 2021 provided by the Korea Disease Control and Prevention Agency (KDCA). The data on the number of people vaccinated per day, type of vaccine, and age of the person who were vaccinated from 26 February to 8 October 2021 are available in [9].

### 4.2 Mathematical model

The model we develop is an extension of the models in [7, 8] to include age-specific transmission rates of the Delta and non-Delta variants. The dynamics of infection with the Delta or non-Delta variants follows an SEIQR scheme, where the subscripts *i* and *v* refer to age group and vaccination, respectively, and the superscripts *nonδ* or *δ* refer to whether the infection is with the non-Delta or Delta variant, respectively. Eight age groups are considered: 0 to 17 are referred to as group 1, 18 to 29 as group 2, and those aged 30 to 39, 40 to 49, and so on until 80 and above, are groups 3 to 8. The model diagram depicted in Figure 1 shows that susceptible (*S*) and vaccinated individuals may be exposed to non-Delta or Delta variants (*E*), with a force of infection *λ*, and become infectious (*I*). Once confirmed, these individuals are isolated and categorized as mild (*Q*^*m*^) or severe (*Q*^*s*^), and eventually recover (*R*) or die. Compartments for individuals who are vaccinated effectively (*V*), ineffectively (*U*), and then developed partial (*P*^*part*^) or full immunity (*P*^*f ull*^) are also considered.

**Figure 1:**
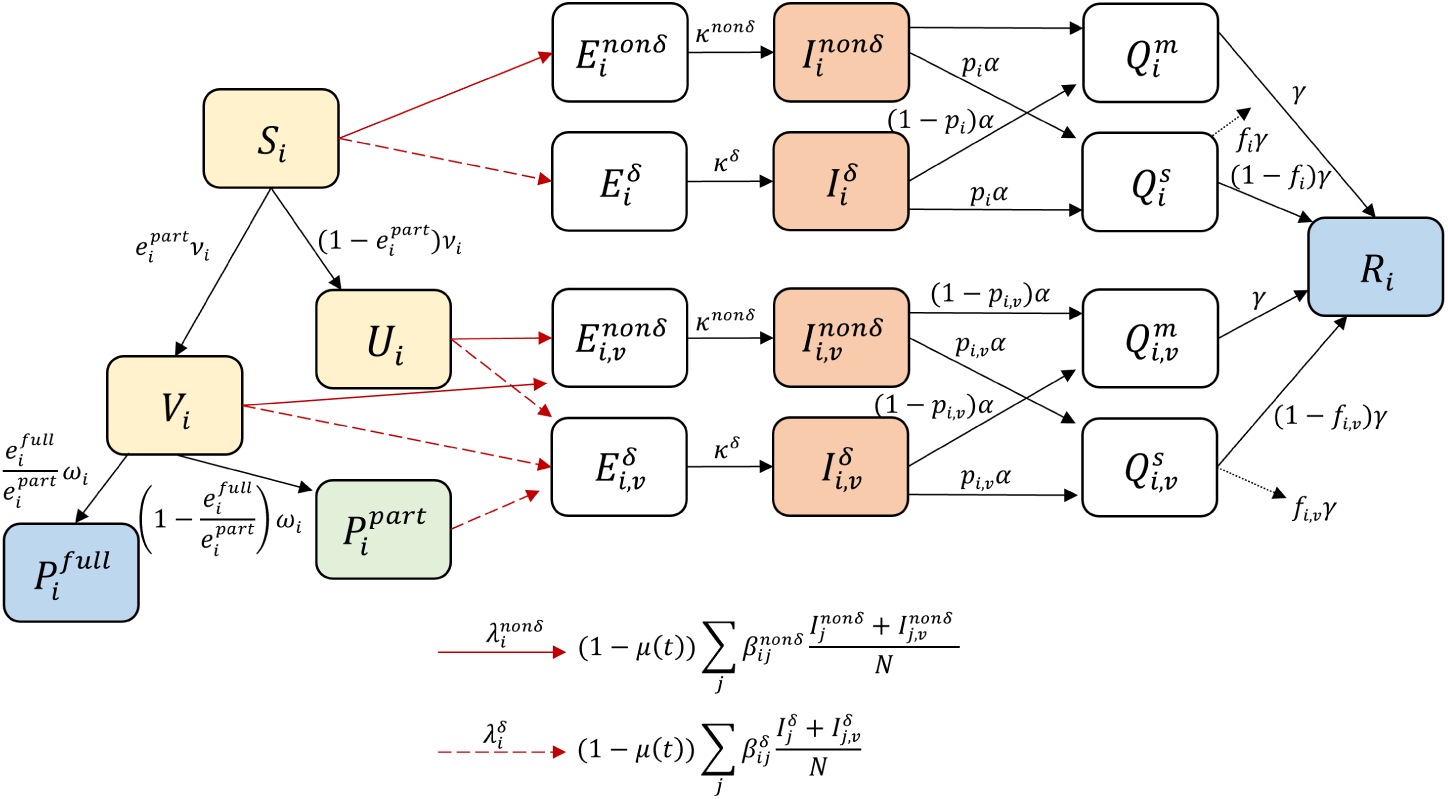
The model flowchart describing the transmission of COVID-19 with the non-Delta or Delta variants. The susceptible class of age group *i*, denoted by *S*_*i*_, can be exposed to the non-Delta or Delta variants 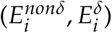 with forces of infection 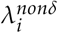 or 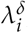, respectively. The transmission reduction factor *µ*(*t*) quantifies the NPIs according to government policy. Also, *S*_*i*_ can become effectively *V*_*i*_ or ineffectively *U*_*i*_ vaccinated, depending on the daily number of vaccinated individuals *ν*_*i*_, and vaccine effectiveness to the non-Delta variant 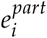. Individuals in *V*_*i*_ become partially 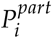 or fully protected 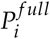 from infection after 1/*ω*_*i*_ days on average, or may be exposed to the non-Delta or Delta variant 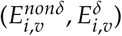. Vaccine effectiveness to the Delta variant is denoted by 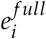. The mean latent period of the non-Delta and Delta variants are 1/*κ*^*nonδ*^ and 1/*κ*^*δ*^ days, respectively. The mean infectious period is 1/*α* days. Those in the infectious classes (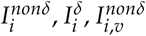 and 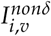) are isolated as soon as confirmed, and are classified as mild 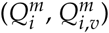 or severe 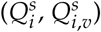. Individuals in the isolated compartments may die or recover (*R*_*i*_) after 1/*γ* days on average. The parameters *p*_*i*_, *p*_*i,v*_ represent the proportion that becomes severe and *f*_*i*_, *f*_*i,v*_ are the mean fatality rates of the unvaccinated and vaccinated individuals, respectively.

The age-specific, vaccine-dependent parameters *e*^*part*^, *e*^*f ull*^, and *ω* consider the proportion of an age group vaccinated with either ChAdOx1 or BNT162b2 [10], effectiveness of these vaccines to the Delta and non-Delta variants [3], and interval between doses [11]. The transmission rates *β* for the Delta and non-Delta variants are estimated using MLE. The transmission reduction factor *µ*(*t*), which quantifies the NPIs according to government policy, is also estimated. The values of *ν, p* and *f* are derived from data. Details of the model development are found in Appendix A.

We apply MLE to estimate the transmission rates of the Delta and non-Delta variants between age groups by considering two periods. Transmission rates of the non-Delta variant were estimated from the period 26 February to 30 June 2021, while the data from 1 August to 10 September, when the proportion of cases with the Delta variant was over 80%, were used to estimate the transmission rates for the Delta variant [12].

### 4.3 Modelling scenarios

To examine the impact of vaccination rollout, we extended the model simulation until 30 December 2021 and analyzed the effect of easing NPIs at different times. Considering that approximately 77% of the population already have at least one dose, we assumed that 85% of the population is vaccinated by 30 November 2021. We consider three dates for the initiation of eased NPIs: 8 October, 18 October, and 1 November. For each date, we simulate three scenarios with different values of the transmission reduction factor *µ*(*t*): *µ* = 0.69 in scenario 1 (S1), *µ* = 0.64 in scenario 2 (S2), and *µ* = 0.52 in scenario 3 (S3). The estimated value obtained from 16 April to 3 June 2021 is S1, and the minimum value estimated in this study is S2 (from 16 June to 11 July 2021). The value of *µ* in S3 was estimated from the third epidemic wave in 2020, when it was SD1, and is a relatively lower value compared to S2 [13].

We also proceeded with simulation-based experiment to study the effects of varying the speed of vaccination, arrival timing of the Delta variant, and intensity of NPIs. In this case, the initial condition of simulations is identical to 26 February 2021 in Korea and the simulation time is set to 365 days. The total number of vaccinated people is assumed to be 40 million, which is approximately 78% of the population in Korea, and the number of people vaccinated daily is set to 150 000, 200 000, or 400 000. Moreover, we fix the order of vaccination from oldest to youngest, that is, 8-7-6-5-4-3-2, excluding group 1. We denote by *t*_*δ*_ the day when the individuals exposed to the Delta variant arrived in the local community, and we set its range from zero to 80 days. Note that in Korea, the date of arrival of the Delta variant corresponds to *t*_*δ*_ = 52. We consider values of *µ* from 0.52 to 0.78 in 0.001 increments. In total, there are 21 141 simulations for the different values of *ν*_*i*_, *t*_*δ*_, and *µ*.

## 5 Results

### 5.1 Parameter Estimation

The estimated transmission rates obtained using MLE form a matrix according to the age groups. The transmission rate matrices, visualized in Figure 2, represent the transmission patterns of the non-Delta (a) and Delta variants (b) in Korea.

**Figure 2:**
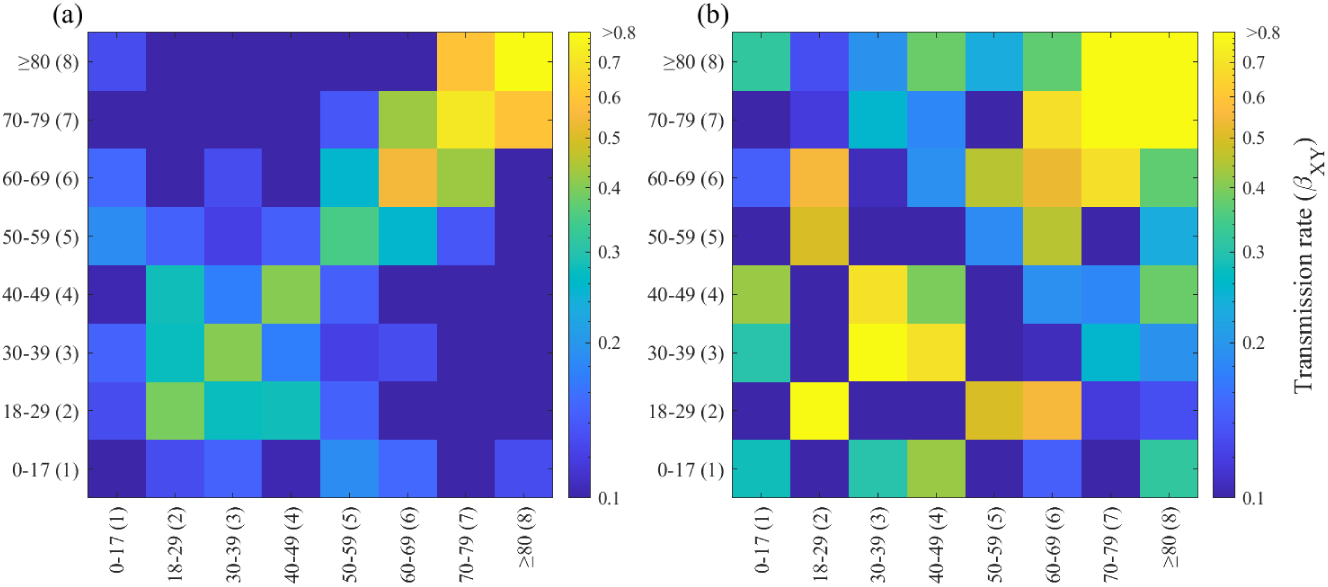
Estimated transmission rates among age groups (a) non-Delta variants from 26 February to 30 June 2021 and (b) Delta variant from 1 August to 10 September 2021.

Since the various government policies including SD implemented during the periods considered in estimating the transmission rates, we normalize the transmission rate matrices to exclude phase-dependent factors. The details of the computation are found in Appendix C.

We use the normalized transmission rate matrices in estimating the factor *µ*(*t*), which represents the reduction in transmission induced by NPIs according to government policy. Table 1 presents the estimates for each phase from 26 February to 8 October 2021. Figure 3 (a) shows the plots of the estimates for the reduction factor *µ*(*t*) (red curve) with the effective reproductive number (ℛ_*t*_, blue curve) described in Appendix C. At the start of the simulation period, Seoul Capital Area was at SD2. When *µ*(*t*) increased from 0.64 to 0.69 around 16 April 2021, ℛ_*t*_ decreased to 0.97 and stayed below 1 until around 16 June. From 16 June to 11 July, *µ* was estimated at 0.64 and ℛ_*t*_ jumped to as high as 1.52. The exponential rise in the proportion of cases infected with the Delta variant (black solid curve in Figure 3 (a)) and daily confirmed cases (Figure 3 (b)) was also observed on the same period, which prompted the start of the fourth wave. On 12 July, the Korean government raised the SD level in Seoul Capital Area to SD4. A steep increase in the cumulative confirmed cases was seen beginning 12 July (Figure 3 (c)), and the estimates for *µ* were the highest from this period until 06 September. At the same time, the proportion of hosts having full immunity (black dashed curve in Figure 3 (a)) increased steadily, reaching up to about 50% by the end of the estimation period. On the last phase, the estimated value of *µ* was 0.70.

**Table 1:** Estimates for the phase-dependent, transmission reduction factor *µ*(*t*), social distancing (SD) level, and range of effective reproductive number ℛ_**t**_ from 26 February to 8 October 2021.

| Phase | Period | Value | SD Level | $\mathcal{R}_t$ |
| --- | --- | --- | --- | --- |
| 1 | 26 Feb to 25 Mar | 0.64 | 2 | [1.13, 1.14] |
| 2 | 26 Mar to 15 Apr | 0.65 | 2 | [1.11, 1.12] |
| 3 | 16 Apr to 3 Jun | 0.69 | 2 | [0.96, 0.97] |
| 4 | 4 Jun to 15 Jun | 0.70 | 2 | [0.94, 0.96] |
| 5 | 16 Jun to 11 Jul | 0.64 | 2 | [1.16, 1.52] |
| 6 | 12 Jul to 25 Jul | 0.76 | 4 | [1.03, 1.09] |
| 7 | 26 Jul to 8 Aug | 0.77 | 4 | [1.07, 1.07] |
| 8 | 9 Aug to 5 Sep | 0.78 | 4 | [0.91, 1.00] |
| 9 | 6 Sep to 19 Sep | 0.71 | 4 | [1.11, 1.21] |
| 10 | 20 Sep to 8 Oct | 0.70 | 4 | [0.94, 1.10] |

**Figure 3:**
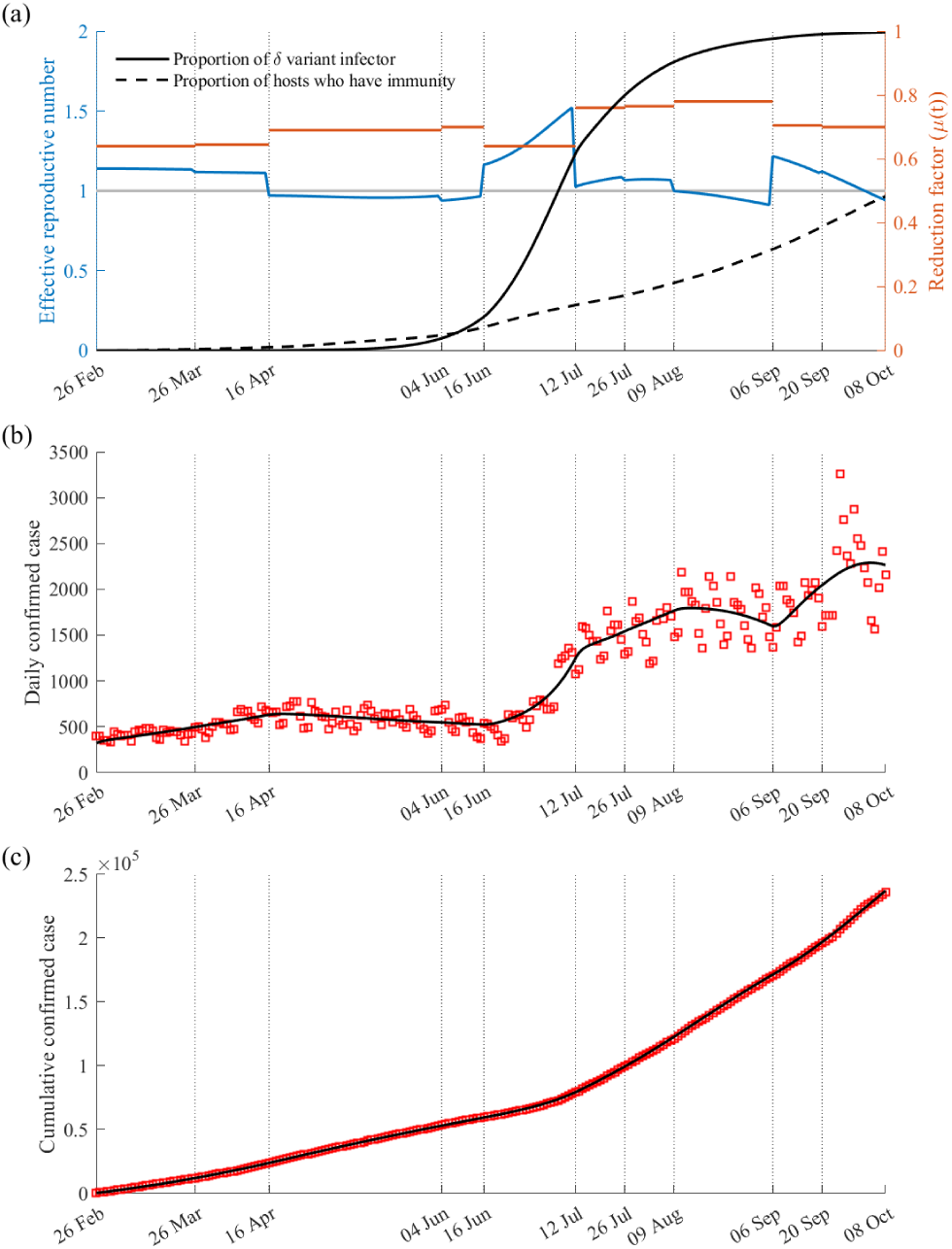
The parameter estimation results obtained by fitting the model to the cumulative confirmed cases. Panel (a) shows the effective reproductive number (blue), estimates of the transmission reduction factor *µ*(*t*) (red), proportion of the active cases infected with the Delta variant (black solid curve), and proportion of the total population who has immunity (black dashed curve). Panel (b) shows the best fit of the model to the daily confirmed cases and (c) the best fit of the model to the cumulative confirmed cases. The boxes represent the data for the daily and cumulative cases from 26 February to 8 October 2021.

To illustrate how *µ* can be used to suggest appropriate SD level, assume that the basic reproductive number of the disease is five, Delta and non-Delta variants can infect the population, 80% of population is vaccinated, and vaccine effectiveness is 62.5%. Then without NPIs, ℛ_*t*_ = 5 × 0.8 × 0.625 = 2.5. To maintain an ℛ_*t*_ below one, SD2 is appropriate since the minimum estimated value of *µ*(*t*) on SD2 was 0.64 and this translates to ℛ_*t*_ = (1 − 0.64) × 2.5 = 0.9. On the other hand if *µ* = 0.52, which represents worst case during SD1, then ℛ_*t*_ = (1 − 0.52) × 2.5 = 1.2. In this case, SD4 may not necessary and can aggravate present economic problems.

In Figure 4, we present the best fit of the model to the data on daily and cumulative cases per age group. The rise in the number of cases in the period from 16 June 2021 was observed across all age groups, with age 18-29 having the most and the elderly groups having the least increase. The model shows a peak (459 cases) on 16 August and another peak (519 cases) on 28 September in age 18-29. Towards the end of the simulation period, a decreasing trend in the number of cases was observed in most age groups except age 0-17, not vaccinated.

**Figure 4:**
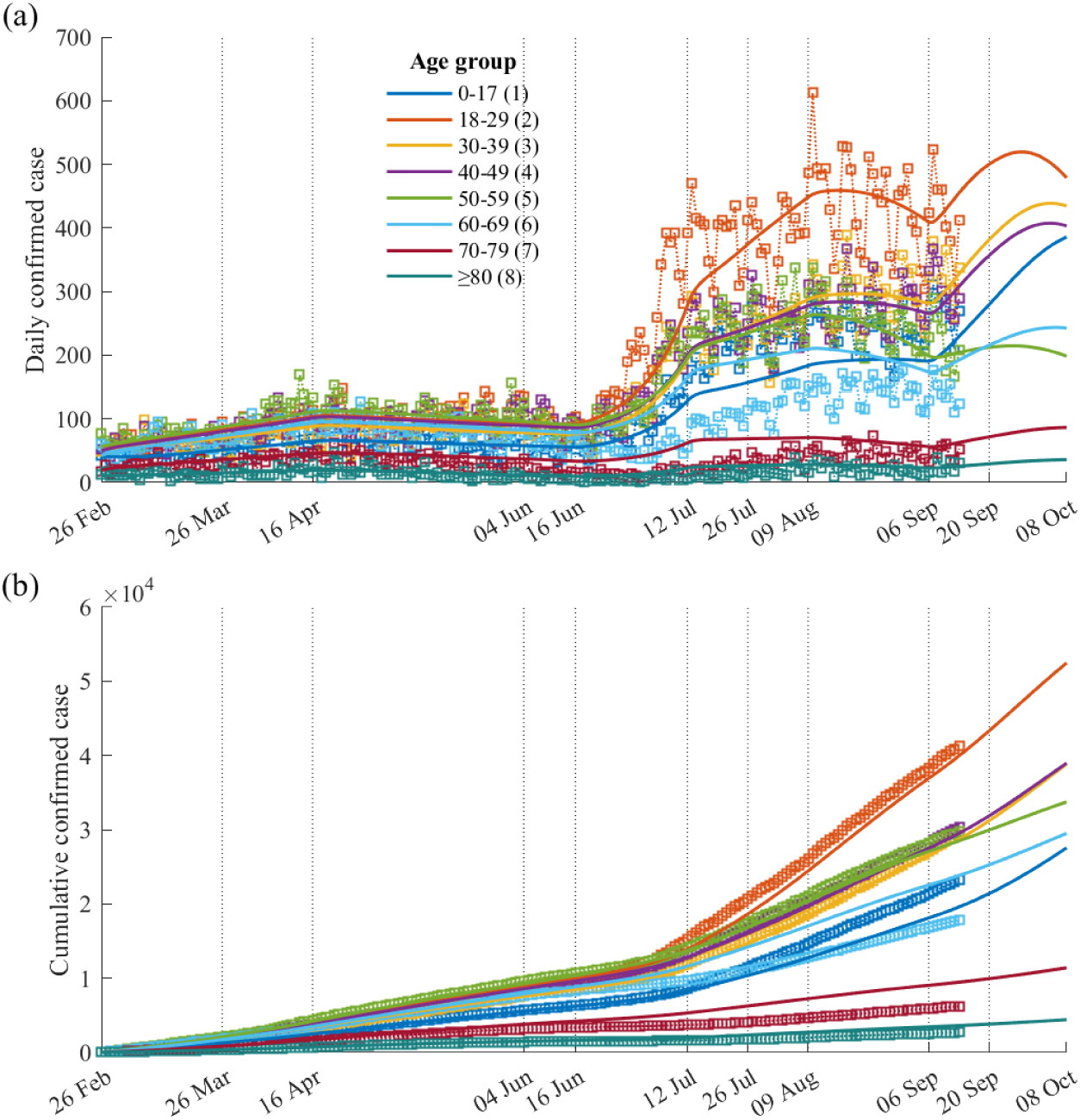
The best fit of the model to the (a) daily confirmed cases and (b) cumulative confirmed cases per age group. The boxes represent the data per age group from 26 February to 10 September 2021.

### 5.2 Analysis on the timing of easing the NPIs

Nine scenarios considering different dates of initiation and levels of eased NPIs are displayed in Figure 5. The red, black, and blue curves, correspond to starting the eased NPIs on 8 October, 18 October, and 1 November 2021, respectively. The solid curves represent S1, dashed curves S2, and dotted curves S3. Panel (a) shows ℛ_*t*_, (b) the daily confirmed cases, (c) the number of severe patients requiring hospital beds, and (d) the cumulative death. The dashed grey curve in panel (c) marks the maximum number of hospital beds (1067) for severe patients in Korea [14]. Results show that the worst case scenario is S3 with 8 October as easing time, wherein the peak number of daily cases, severe patients, and cumulative deaths could reach 8207, 1617, and 6528, respectively. But if the easing to S3 was started later, on 18 October or 1 November, the peak numbers could be reduced to 4385 (40.48% reduction) or 1961 (76.11% reduction) for the peak daily cases, 879 (45.64% reduction) or no increase for the peak number of severe patients, and 3847 (42.07% reduction) or 2313 (64.57% reduction) for the cumulative deaths, compared to when the easing was initiated earlier. The rest of the simulations show a decreasing trend of daily confirmed cases and number of severe patients. The simulations illustrate the importance of the timing of easing NPIs in preventing the occurrence of another epidemic wave. All scenarios, except the worst case, show that the number of severe patients did not reach the limit (1067) of Korea.

**Figure 5:**
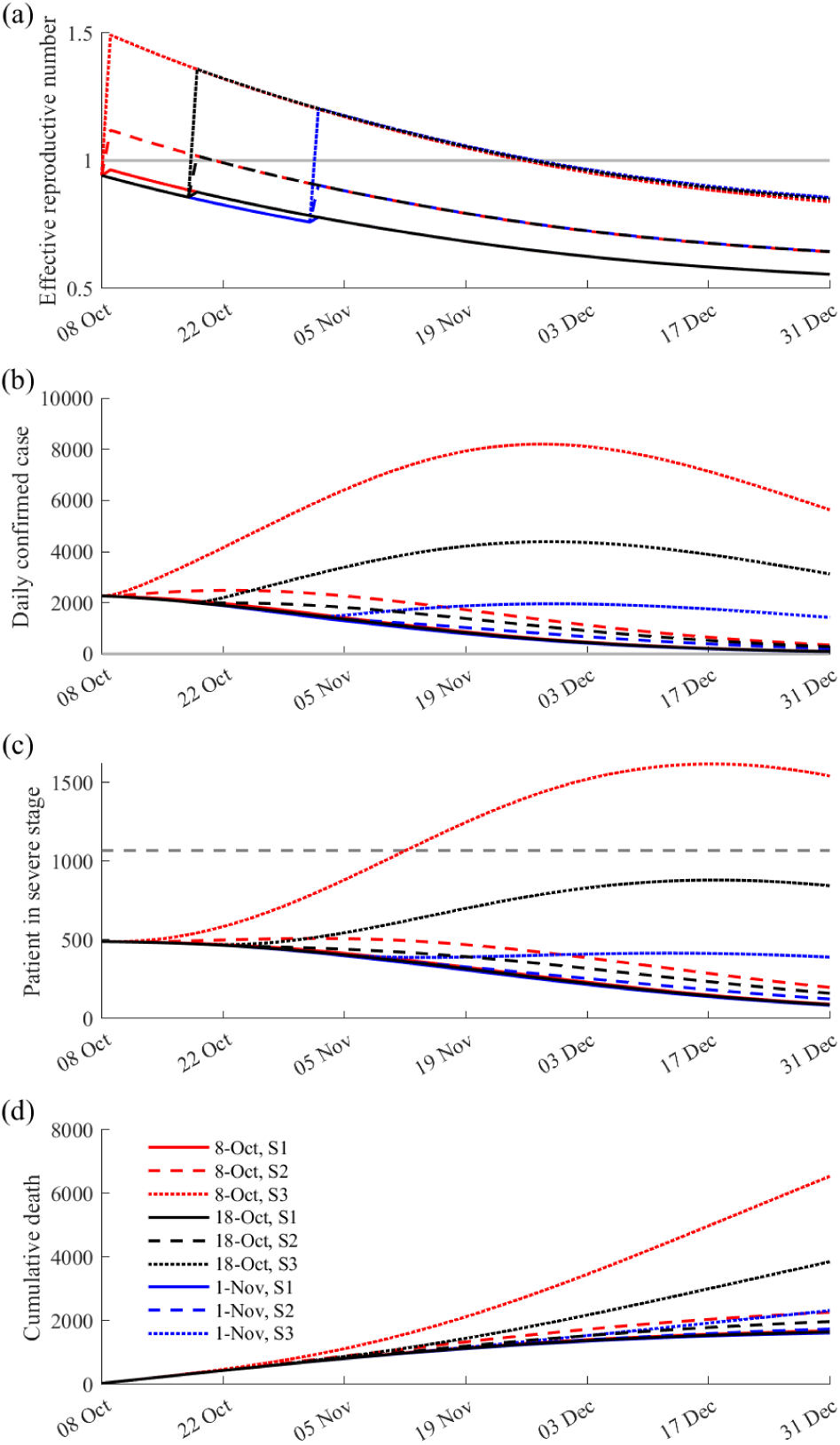
Results on the effect of easing NPIs at different times. Panel (a) shows the effective reproductive number, (b) number of daily confirmed cases, (c) number of severe patients requiring hospital beds, and (d) cumulative number of deaths. Colours and shapes of curves indicate the timing of initiation (8 October, 18 October, and 1 November) and level of eased NPIs (S1, S2, and S3). Grey dashed line in (c) indicates the number of available beds (1067) for severe patients in Korea.

### 5.3 Analysis on the vaccination speed, NPIs, and arrival timing of the Delta variant

Figure 6 displays the simulation results when *µ* = 0.67, *t*_*δ*_ = 52, and *ν*_*i*_ = 200 000 or 400 000. In panel (a), ℛ_*t*_ and the proportion of cases infected with the Delta variant are displayed. The magenta asterisk indicates *t*_*δ*_ = 52. When *ν*_*i*_ = 200 000, ℛ_*t*_ increased up to 1.4, while if *ν*_*i*_ = 400 000, ℛ_*t*_ decreased to below one. In panels (b) and (c), a second peak with more than 2000 cases and 600 severe patients is observed when *ν*_*i*_ = 200 000 (red curve), whereas no second peak is observed when *ν*_*i*_ = 400 000 (blue curve). Panel (d) shows that the cumulative deaths when *ν*_*i*_ = 200 000 can reach 4.48 times higher compared to when *ν*_*i*_ = 400 000. Panels (e) and (f) display the cumulative number of cases and deaths of each age group, with 34% of the cases occurring in age group 2 and 68% of the deaths occurring in age group 8 when *ν*_*i*_ = 200 000. These results highlight the effect of the daily number of vaccination to the occurrence of another epidemic wave.

**Figure 6:**
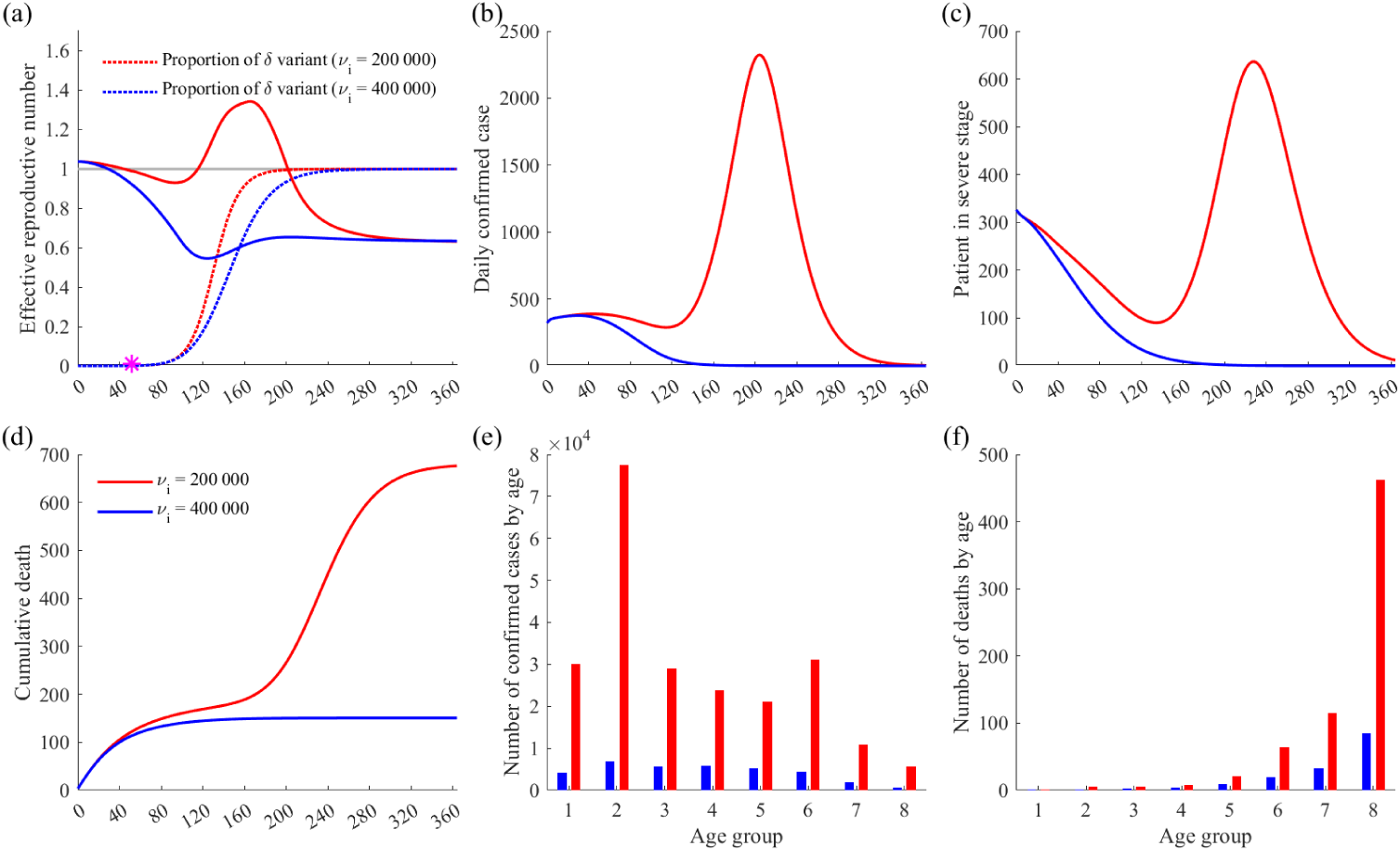
The simulation result with *µ* = 0.67, *t*_*δ*_ = 52, and daily number of vaccination set to 200 000 or 400 000. Panel (a) shows the effective reproductive number, (b) number of daily confirmed cases, (c) number of severe patients requiring hospital beds, (d) cumulative number of deaths, (e) age-dependent number of confirmed cases, and (f) age-dependent number of deaths. Red and blue colours indicate that the number of daily administered vaccines is 200 000 and 400 000, respectively.

Figure 7 shows a heat map summarizing the results of the 21 141 simulations. The columns represent the daily vaccination number (*ν*_*i*_ = 150 000, 200 000, and 400 000) and the rows correspond to the number of confirmed cases, deaths, and peak number of severe patients requiring hospital beds. In each panel, the *x* axis represents *t*_*δ*_ and the *y* axis corresponds to *µ*. The dotted red curve in panels (g), (h), and (i) marks the maximum number of hospital beds (1067) for severe patients in Korea.

**Figure 7:**
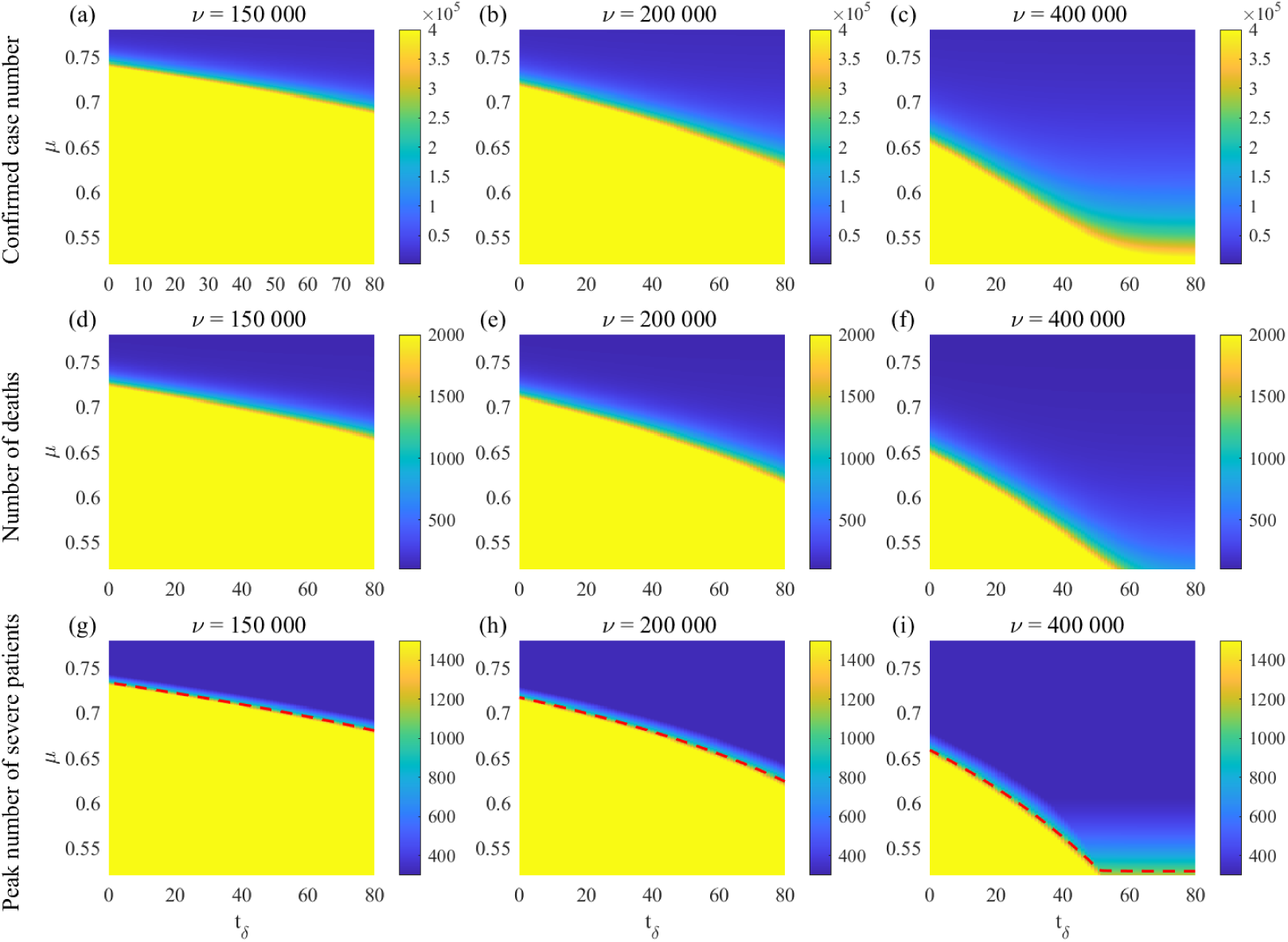
The simulation results of the 21,141 runs. The columns from left to right indicate the number of daily administered vaccines as 150 000, 200 000, and 400 000, respectively. Panels (a), (b), and (c) show the number of confirmed cases; (d),(e), and (f) the number of deaths; and (g),(h), and (i) the peak number of severe patients requiring hospital beds. Note that red dotted curve in panels (g),(h), and (i) indicates the maximum number of available beds (1067) for severe patients in Korea.

## 6 Discussion

The transmission rate matrix in this study was apparently different between the Delta and non-Delta variants and by age groups. The transmission rate with non-Delta variant was high between the same age groups. However, transmission with the Delta variant was much higher among age groups of 18-29 and 30-39, and an increased transmission was observed between those age groups and age groups of 50-59 and 60-69. This matrix reflects the distinct epidemiologic characteristic of the COVID-19 epidemic after Delta became the dominant variant of SARS-CoV-2 in Korea: 1) incidence in younger age groups, those in 20s and 30s, increased; 2) and the major source of transmission moved from group-related or institutional outbreaks to individual contacts [15, 16]. These results indicate an increased transmission with Delta variant among household contacts, which is coherent with previous studies [17, 18].

Higher transmission rates with the Delta variant compared to the non-Delta variant were also observed in the elderly groups, which may have resulted from breakthrough infections in nursing homes and assisted living facilities [27]. With the absence of vaccination, a much higher transmission rate with the Delta variant was expected, but was not observed in the youngest age group of 17 years old and below. This phenomenon was probably a consequence of the Korean government policy on limiting the number of attendance in school to two-thirds since October 2020 [19]. According to the data, among all the COVID-19 cases of those aged 17 years old and below in Korea, only about 15% are infected from schools during this period [20].

We also analyzed the reduction in transmission induced by NPIs according to the government policy, presented by *µ*(*t*). SD2 and SD4 in Korea corresponded to *µ*(*t*) values from 0.64 to 0.69, and from 0.70 to 0.78, respectively. Values of *µ*(*t*) close to 0.7 may represent SD3 in Korea. Considering the basic reproductive number of a disease, vaccine effectiveness, and the amount of administered vaccines, the fitted values of *µ*(*t*) can be an effective measures of the government policy. This parameter would be informative to the healthcare professionals and policy makers on designing prevention and control measures for emerging or reemerging infectious diseases.

Considering both the medical and economic impact of NPIs, the Korean government plans to reduce SD, termed ‘Back to normal or With-Corona’, on 1 November 2021 [14]. Because vaccination and immunization takes time, we explored the effect of delaying the initiation of eased SD in Figure 5. When SD was eased to a minimal value (S3), no large-scale re-emergence of infections was observed when S3 was delayed to 1 November, as opposed to when it was initiated on 8 or 18 October. In other words, gradually easing NPIs might be necessary when implementation is earlier. For example, in Korea, the maximum number of people allowed in a private gathering after 6:00PM became four (ten for fully vaccinated) from two since 18 October, although SD remained at SD4 [21].

In our generalized simulation on the effects of vaccination and NPIs, the larger reduction of transmission factor (*µ*(*t*)) and faster vaccination rollout or earlier implementation of stricter NPIs (e.g., expanding genomic testing to people entering from high-risk countries or elevating SD) resulted in less incidence, severe cases, and deaths. We are aware that the supply of vaccines is not enough globally, but fast administration of vaccines can block the occurrence of another wave. In other words, epidemic can be controllable and stable if the vaccination speed is faster.

There are several limitations of this research. First, the contact pattern in the matrices was already affected by NPIs and might be not identical to the contact pattern before COVID-19, necessitating the normalization done on the matrices. For example, transmission rate among those age 0-17 might be underestimated because of restricted attendance in schools. Second, vaccine waning is not considered. However, booster shots are administered and vaccine hesitancy remains low in Korea, which mean that the effect of administered vaccines might be maintained [22]. Third, our age-age matrix focused on the Delta variant, and we were not able to examine transmission of other previous variants. The coverage of genomic surveillance among COVID-19 cases in Korea has steadily increased, and it covers about 30% of all confirmed cases. Therefore, the number of cases with genomic results before the occurrence of the Delta variant was small to be analyzed separately by variant types. Our result highlighted the transmission pattern of Delta, which is currently the dominant variant in most countries.

## 7 Conclusion

This study illustrated the higher transmissibility of the Delta relative to the non-Delta variant among various age groups in Korea by developing the transmission rate matrix based on MLE. The method used to quantify the effects of the vaccination and NPIs simultaneously. The model simulation results emphasize the importance of simultaneously applying interventions, such as SD, screening measures at the entry points, and vaccination. Another epidemic wave can be avoided if a strict SD policy is not relaxed too early. Transmission rate of the Delta variant of COVID-19 is high and oral treatment has not yet been provided widely, but the epidemic is still ongoing, so we suggest that NPIs are still necessary to control the epidemic and reduce the number of severe cases to prevent a burden to the healthcare system. Even with the same SD level, model simulations showed that outcomes may change depending on vaccine administration and emergence of a highly transmissible variant. If vaccination is slow, then NPIs should be strictly implemented to lessen the medical and socio-economic burden of COVID-19 to the society.

## Data Availability

All data produced are available online at

http://ncov.mohw.go.kr/

## 8 Ethics approval

The study was conducted according to the guidelines of the Declaration of Helsinki, and approved by the Institutional Review Board of Konkuk University (7001355-202101-E-130)

## Appendix A

### Development of the mathematical model

Susceptible individuals in age group *i*, denoted by *S*_*i*_ (*i* = 1, 2, …, 8), who are unvaccinated become exposed to the non-Delta 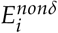 or Delta 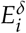 variants with forces of infection 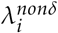 or 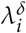, respectively. These exposed individuals become infectious 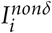 after 1/*κ*^*nonδ*^ or 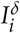 after 1/*κ*^*δ*^days. Here we assume that the latent period of infection with the Delta variant is shorter than the non-Delta variant [23, 24].

To account for vaccination, individuals in *S*_*i*_ can become effectively *V*_*i*_ or ineffectively *U*_*i*_ vaccinated, depending on the daily number of vaccinated individuals in age group *i*, denoted by *ν*_*i*_, and vaccine effectiveness to the non-Delta variant 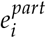. Individuals in *V*_*i*_ eventually become partially 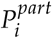 or fully protected/immune 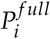 from infection after 1/*ω*_*i*_ days. Fully protected individuals are assumed to be immune to COVID-19, while partially protected individuals can only be infected with the Delta variant. Vaccinated individuals who are exposed to the non-Delta 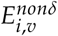 or Delta 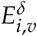 variant eventually progress to the vaccinated infectious classes 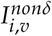 or 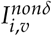.

We assume that individuals in the infectious classes (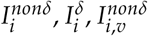 and 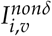) are isolated as soon as they were confirmed, and are classified as mild 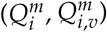 or severe 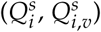. The mean infectious period of an infected individual is 1/*α* days, and *p*_*i*_ or *p*_*i,v*_ denote the proportions that an infected unvaccinated or vaccinated individual progresses to a severe case. Individuals in the isolated compartments recover 1/*γ* days on average and the mean fatality rates of the unvaccinated and vaccinated groups are denoted by *f*_*i*_ and *f*_*i,v*_, respectively.

The following system of ordinary differential equations describes the developed age-structured model:

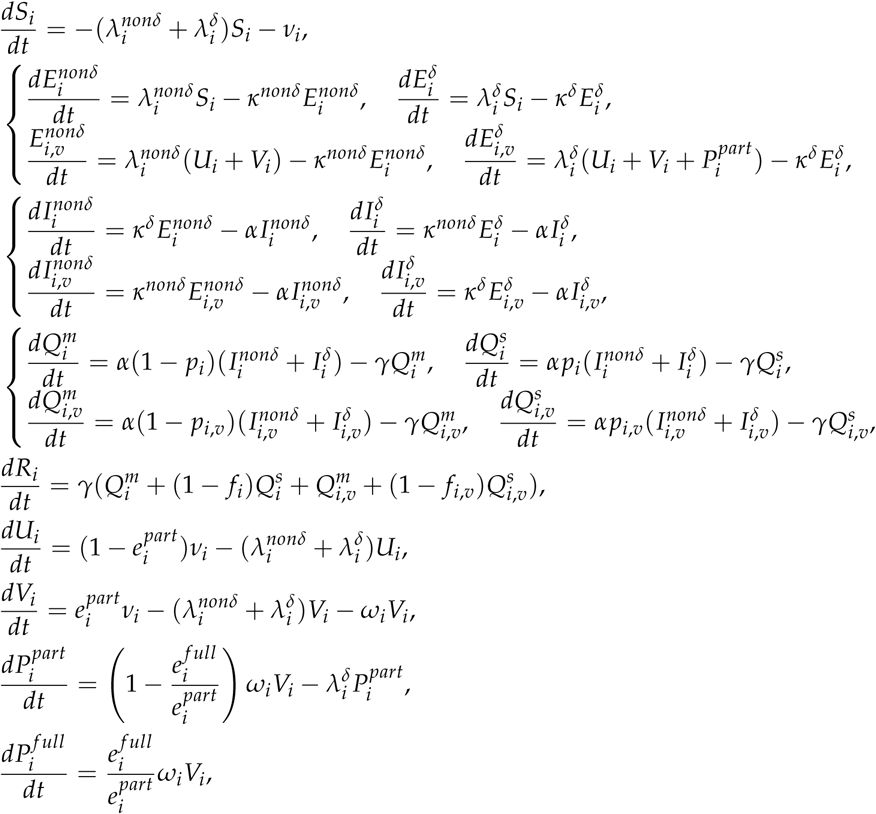

where the forces of infection 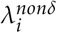 and 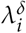 are defined as

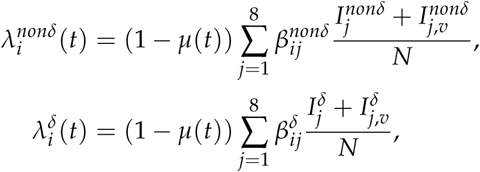

*µ*(*t*) is the phase-dependent, transmission reduction factor accounting for NPIs and its estimated values are in Table 1, 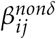 and 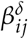 are the transmission rates between age groups, and

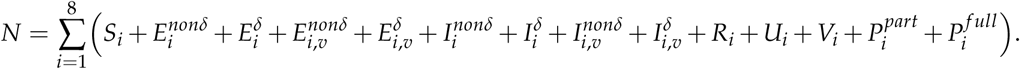

Tables 2 and 3 summarize the age-specific and non-age-specific model parameters and their values.

**Table 2:** Non-age-specific parameters of the model.

| Symbol | Description | Value | Reference |
| --- | --- | --- | --- |
| $1/\kappa^{non\delta}$ | Mean latent period of the non-Delta variant | 4 days | [23, 24] |
| $1/\kappa^{\delta}$ | Mean latent period of the Delta variant | 2 days | [23, 24] |
| $1/\alpha$ | Mean infectious period | 6 days | [24, 25] |
| $1/\gamma$ | Mean duration of isolation (hospitalization) | 20 days | [26] |

**Table 3:** Age-specific parameters of the model calculated using data from [10, 11] and vaccine effectiveness summarized in Table 4.

| Symbol | Description | Age group |  |  |  |  |  |  |  |
| --- | --- | --- | --- | --- | --- | --- | --- | --- | --- |
|  |  | 1 | 2 | 3 | 4 | 5 | 6 | 7 | 8 |
| $p_i$ | Proportion of unvaccinated group $i$ that becomes severe | 0 | 0 | 0.01 | 0.02 | 0.04 | 0.07 | 0.13 | 0.26 |
| $p_{i,v}$ | Proportion of vaccinated group $i$ that becomes severe | 0 | 0 | 0 | 0.01 | 0.01 | 0.03 | 0.02 | 0.05 |
| $f_i$ | Mean fatality rate of unvaccinated group $i$ (1/day) | 0 | 0 | 0 | 0 | 0 | 0.01 | 0.03 | 0.13 |
| $f_{i,v}$ | Mean fatality rate of vaccinated group $i$ (1/day) | 0 | 0 | 0 | 0 | 0 | 0 | 0 | 0.01 |
| $e_i^{part}$ | Vaccine effectiveness to non-Delta | 0.94 | 0.93 | 0.91 | 0.9 | 0.91 | 0.76 | 0.83 | 0.92 |
| $e_i^{full}$ | Vaccine effectiveness to Delta | 0.88 | 0.87 | 0.85 | 0.84 | 0.85 | 0.69 | 0.76 | 0.86 |
| $1/\omega_i$ | Mean interval from first dose to immunity (1/day) | 35.44 | 36.4 | 40.12 | 41.91 | 39.44 | 64.08 | 52.52 | 38.95 |

Table 4 shows the vaccine effectiveness of ChAdOx1 and BNT162b2 to the non-Delta and Delta variants after the first and second doses. Table 5 shows the proportion of each age group vaccinated with ChAdOx1, and the calculated values of 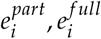 and *ω*_*i*_. The average duration from the first dose to having immunity (1/*ω*_*i*_) and vaccine effectiveness to the non-Delta and Delta variants 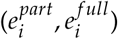 of each age group are calculated depending on the vaccine effectiveness and type of vaccine given to a proportion of an age group. We assume that the interval between doses of ChAdOx1 and BNT152b2 are 11 and 4 weeks, respectively.

**Table 4:** Vaccine effectiveness of ChAdOx1 and BNT162b2 to the non-Delta and Delta variants depending on the number of doses [3].

|  | ChAdOx1 |  | BNT162b2 |  |
| --- | --- | --- | --- | --- |
|  | First dose | Second dose | First dose | Second dose |
| Vaccine effectiveness to the non-Delta variant | 0.49 | 0.74 | 0.47 | 0.94 |
| Vaccine effectiveness to the Delta variant | 0.30 | 0.67 | 0.36 | 0.88 |

**Table 5:** Proportion of age group *i* administered with ChAdOx1 from data [10] and the calculated age-dependent vaccine effectiveness to the Delta and non-Delta variants, and average duration to develop full immunity.

| Description | Age group |  |  |  |  |  |  |  |
| --- | --- | --- | --- | --- | --- | --- | --- | --- |
|  | 1 | 2 | 3 | 4 | 5 | 6 | 7 | 8 |
| Proportion of age group $i$ administered with ChAdOx1 | 0.00 | 0.03 | 0.15 | 0.21 | 0.13 | 0.93 | 0.55 | 0.11 |
| $e_i^{part}$ | 0.94 | 0.93 | 0.91 | 0.9 | 0.91 | 0.76 | 0.83 | 0.92 |
| $e_i^{full}$ | 0.88 | 0.87 | 0.85 | 0.84 | 0.85 | 0.69 | 0.76 | 0.86 |
| $1/\omega_i$ (days) | 35.44 | 36.40 | 40.12 | 41.91 | 39.44 | 64.08 | 52.52 | 38.95 |

## Appendix B

### MLE formulation

To establish the likelihood function to be optimized, we first divide each age group into three subgroups to distinguish COVID-19 outcomes: infected (Λ_*Ii*_), unvaccinated and uninfected (Λ_*Si*_), and effectively vaccinated and uninfected (Λ_*Vi*_), where *i* indicates the age group. Let *β*_*XY*_ be the transmission rate from age group *Y* to *X*. Assuming a homogeneous mixing of the population, and that the transmission events are exponentially distributed, the probability that an individual *x* in subgroup Λ at the time *t* − 1 is still uninfected until the next time *t* is given by

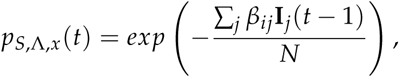

where **I**_*j*_(*t* − 1) denotes the number of hosts in group *j* which can spread the disease at time *t* − 1, and *N* is the total population. We can set *N* as a constant because the number of isolated individuals (or deceased) is less than 0.02% of the population. On the other hand, the probability that an individual *x* in group Λ is uninfected at the time *t* − 1 but becomes infected at *t* is

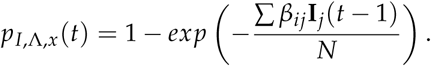

Hence, an individual who stays uninfected until final time *t*_*f*_ has probabilities *p*_*S*,Λ,*x*_(*t*) at each time until *t*_*f*_. In addition, an individual can be vaccinated at time *t*_*Vi,x*_, have immunity after *t*_*e,i*_ days, and stay uninfected until final time *t*_*f*_. An individual infected at time *t*_*Ii,x*_ has probabilities *p*_*S*,Λ,*x*_(*t*) until *t*_*Ii,x*_ − 1 and *p*_*Ii*,Λ,*x*_ (*t*_*Ii,x*_).

The likelihood function *L* is formulated as

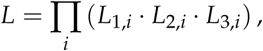

where

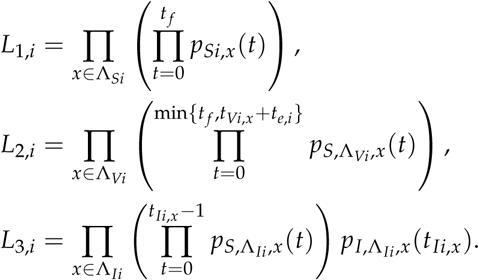

By taking the logarithm of *L* and using the optimization toolbox in MATLAB, we obtain a matrix containing the transmission rates of the Delta and non-Delta variants between age groups.

## Appendix C

### Normalized transmission rate matrices and the effective reproductive number

Let the transmission rate matrices estimated from 26 February to 30 June and from 01 August to 10 September be 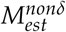 and 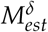, respectively. First, the reproductive number of COVID-19 using 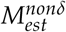 is calculated via the next-generation-matrix method and denoted as 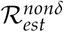. Second, a normalized transmission rate matrix of the non-Delta variant, *M*^*nonδ*^, is formulated as 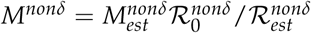, where 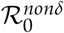, equal to 3.17, is the basic reproductive number of COVID-19 estimated from various studies before Delta variant became the major strain [27]. Third, a normalized transmission rate matrix of the Delta variant is formulated similarly as 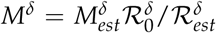, where 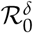 is reproductive number calculated using 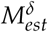 and 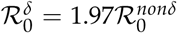 [28]. The normalized transmission rate matrices, *M*^*nonδ*^ and *M*^*δ*^, are used in this research.

The effective reproductive number ℛ_*t*_ is a time-dependent measure of the average number of secondary cases from a single infectious individual [29]. In this study, ℛ_*t*_ is calculated using the next-generation matrix method [30]. Using the normalized transmission rate matrices, ℛ_*t*_ of the non-Delta and Delta variants are 3.17 and 6.24, respectively, if there are no NPIs and behavior change.

